# Frequent Daytime Napping is Detrimental to Human Health: A phenotype-wide Mendelian Randomization Study

**DOI:** 10.1101/2020.01.20.20017723

**Authors:** Lanlan Chen, Aowen Tian, Zhipeng Liu, Miaoran Zhang, Xingchen Pan, Chang He, Wanqing Liu, Peng Chen

## Abstract

**Background:** It remains controversial whether daytime napping is beneficial for human health.

**Objective:** To examine the causal relationship between daytime napping and the risk for various human diseases.

**Design:** Phenotype-wide Mendelian randomization study.

**Setting:** Non-UK Biobank cohorts reported in published genome-wide association studies (GWAS) provided the outcome phenotypes in the discovery stage. The UK Biobank cohort provided the outcome phenotypes in the validation stage.

**Participants:** The UK Biobank GWAS included 361,194 European-ancestry residents in the UK. Non-UKBB GWAS included various numbers of participants.

**Exposure:** Self-reported daytime napping frequency.

**Main outcome measure:** A wide-spectrum of human health outcomes including obesity, major depressive disorder, and high cholesterol.

**Methods:** We examined the causal relationship between daytime napping frequency in the UK Biobank as exposure and a panel of 1,146 health outcomes reported in genome-wide association studies (GWAS), using a two-sample Mendelian randomization analysis. The significant findings were further validated in the UK Biobank health outcomes of 4,203 human traits and diseases. The causal effects were estimated using a fixed-effect inverse variance weighted model. MR-Egger intercept test was applied to detect horizontal pleiotropy, along with Cochran’s Q test to assess heterogeneity among the causal effects of IVs.

**Findings:** There were significant causal relationships between daytime napping frequency and a wide spectrum of human health outcomes. In particular, we validated that frequent daytime napping increased the risks of major depressive disorder, obesity and abnormal lipid profile.

**Interpretation:** The current study showed that frequent daytime napping mainly had adverse impacts on physical and mental health. Cautions should be taken for health recommendations on daytime napping. Further studies are necessary to precisely define the best daytime napping strategies.

## Introduction

As a common habit practiced by people world-wide, daytime napping is widely deemed to be beneficial for human health. However, epidemiological investigations reached controversial conclusions^1^. For instance, some studies have demonstrated that a midday nap could enhance immune system by restoring the neutrophil count to baseline values ^2^, reduce the risk of hypertension and metabolic diseases ^3 4^ and improve alertness, learning capacity and mental health ^5-9^, while others found that daytime napping could increase the risks of diabetes mellitus ^10-12^, hypertension and cardiovascular events ^13 14^, obesity ^15^ and cognitive impairment ^16-18^. One of the main reasons underlying these contradictory conclusions is the difficulty in determining the causal role of daytime napping in human health and disease. Observational studies are known to be readily confounded by factors such as lifestyles, socioeconomic status and other unknown conditions, which may lead to uncertain conclusions ^19 20^.

The Mendelian Randomization (MR) approach has been proposed over the recent years to solve the aforementioned problems ^21 22^, which has been widely used for examining the causal relationship between two phenotypes or traits. MR is a statistical method employing genetic variants (commonly known as simple nucleotide variants or SNVs) as instrumental variables (IVs) to infer the causal effect of a modifiable non-genetic risk factor on the disease outcome of interest ^19^. Up to date, MR has made great achievement in many fields, such as revealing the casual relationship between telomere length and neoplasm ^23^, unveiling the deleterious effects of LDL (low density lipoprotein) cholesterol on type-2 diabetes ^24^, as well as clarifying the causal impact of phospholipase A2 activity on coronary heart disease ^25^. There have been investigations about the causality of daytime napping on particular diseases, e.g. breast cancer and Alzheimer’s disease ^26 27^. However, no MR analysis study has been performed so far to investigate the potentially causal effect of daytime napping on a broad spectrum of human health related traits and diseases. Examining the causal relationship between daytime napping and overall long-term health outcomes is of importance to disease prevention.

Daytime napping is a sleep-related trait for which the inter-individual variation in the length or frequency of napping were demonstrated to be significantly affected by genetic variance ^28^. Sleeping phenotypes including the daytime napping have a moderate to high heritability ^29^. A few previous GWAS have also identified genetic alleles significantly associated with the frequency or the duration of daytime napping ^29 30^. Using the UK Biobank (UKBB) data, a GWAS has also mapped loci associated with napping frequency as a categorical trait ^30^. These studies provide opportunities to use genetic variants as instrumental variables to test the causal relationship between daytime napping and other traits related to human health and disease.

In this study, we report a comprehensive view of the health impact of daytime napping via MR analyses by leveraging the GWAS data from the UKBB and publicly available non-UKBB GWAS data to examine the impact of daytime napping on a wide variety of disease and health traits, aiming to evaluate the causal relationship that could provide appropriate guidance for health management and disease prevention.

## Methods

The current study was consisted of a discovery stage and a validation stage (Figure 1). The discovery stage MR analysis employed daytime napping frequency phenotype in the UK Biobank (UKBB) as exposure and the outcomes in non-UKBB GWAS. The validation stage used the same exposure and the phenotypes in UKBB as outcomes.

**Figure 1.**
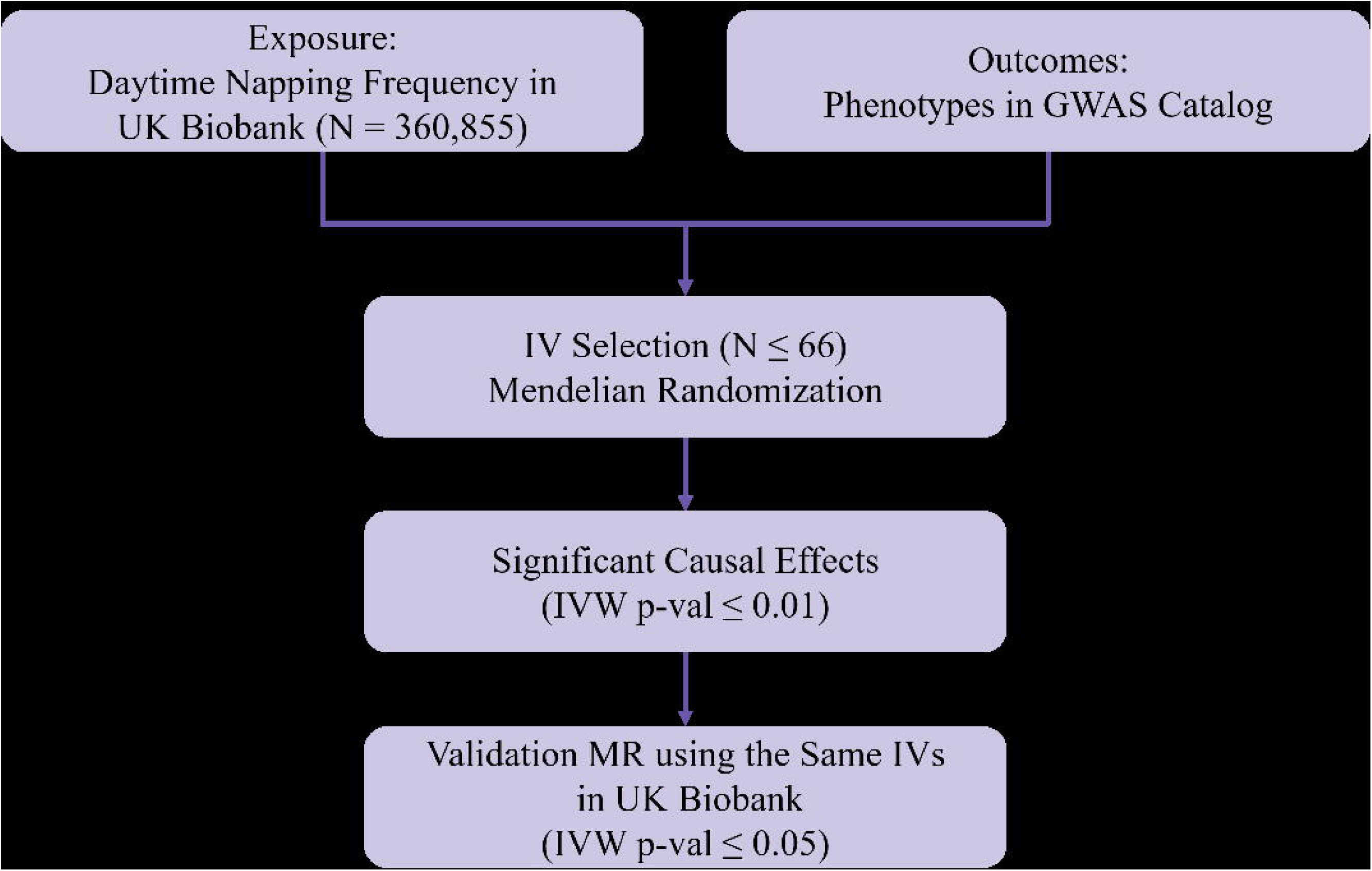
The workflow of our discovery and validation MR analyses.

### Ethics statement

This study used publicly available summary data without accessing to any identifiable information of study subjects or any intervention with living human subjects, thereby is exempt from review by Institutional Review Boards. The study was conducted in line with the principles of Helsinki Declaration.

### Patient and public involvement

It is not possible to involve patients or the public in the design, or conduct, or reporting, or dissemination plans, or manuscript writing of our study. No intervention with living human subjects ever happened in this study.

### UK Biobank phenotype, genetic data and association testing

UK Biobank (UKBB) is a prospective cohort study of UK residents aged 40 through 69 European ancestry. Participants were invited to a recruiting center, where the questionnaires were completed, physical measurements were made, and blood, urine and saliva samples were obtained ^31^. Genome-wide genotypes of 820,000 genomic variants are available.

The GWAS summary statistics used in this study were made by Benjamin Neale’s group (http://www.nealelab.is/uk-biobank/). Briefly, the data QC, genotype imputation and association testing were performed as following. Prior to imputation, multi-allelic SNPs (number of alleles > 2), low frequency SNPs (MAF < 1%), and SNPs appearing to be completely missing in ∼10% in each batch were removed. Array genotypes were phased using SHAPEIT3 and imputed off the merged HRC, UK10K and 1000 Genomes cosmopolitan haplotypes using IMPUTE4 ^32 33^. The imputed 90 million SNPs were filtered to 13.7 million QC SNPs for association testing. The association testing of various phenotypes was conducted using linear or logistic regression model adjusted for age, sex, square age, interaction of sex and age, interaction of sex and square age, and the first 20 PCs. Only samples of European ancestry (N= 361,194) were included in the association tests. Further information on genotype QC, imputation and association testing of UKBB data can be found in the Supplementary Methods.

The daytime napping frequency was categorized by how often the napping was taken (i.e. 1 for never/rarely, 2 for sometimes and 3 for usually), which was available for 360,855 European-ancestry people.

### Summary statistics of non-UKBB GWAS

We searched the GWAS catalog (www.ebi.ac.uk/gwas) and MR base (www.mrbase.org) to include summary statistics from GWAS that were independent of UKBB and those used individuals of European ancestry (i.e. non-UKBB GWAS). For those included UKBB samples in meta-analysis, the summary statistics which were calculated from non-UKBB samples were obtained from the original literatures whenever possible.

### Instrumental variables (IV) selection

We selected SNPs as IVs based on the assumption in principle of MR ^21^. Specifically, we selected IVs with p-value ≤ 5 × 10^−8^, which were further quality-controlled based on MAF (≥ 0.01) and variant-call confidence. We further dropped IVs with Bonferroni corrected association p-value with an outcome ≤0.05 ^34 35^. Qualified IVs were clumped based on their pair-wise linkage disequilibrium (LD, r^2^ < 0.01) and association p-values with the exposure. Data harmonization was carried out to ensure that the effects on the exposure and the outcome were estimated using the same allele (Supplementary Methods).

### Mendelian randomization and statistical analysis

In our MR analysis, the association coefficient of each IV with the outcome was weighted by its association coefficient with the exposure using Wald ratio estimation. The estimates of all valid IVs were combined by an inverse-variance weighted (IVW) meta-analysis ^36^ (Supplementary Methods). We used MR-Egger regression to detect the pleiotropic effect of IVs and Cochran’s Q test for heterogeneity estimation ^37^. We discarded the casual effects with MR-Egger intercept p-value ≤0.05 or Cochran’s Q p-value ≤0.05. For the discovery of significant casual effects, we consider IVW p-value ≤0.01 for our discovery stage and IVW p-value ≤0.05 in our validation stage.

All the data processing and statistical analysis were performed with Perl version 5.9.11 (https://www.perl.org/) and R version 3.4.3 (https://www.r-project.org/). The R package ‘TwoSampleMR’ version 0.4.25 (https://github.com/eleporcu/TWMR) was employed for MR analysis ^38^.

### Sensitivity analysis

In our leave-one-out sensitivity analysis, we excluded each SNP step by step to estimate the remaining effect size, with the aim to detect the IVs dominantly driving the final estimation and to further assess the potency of each SNP ^39 40^. The same statistical methods as in our MR analyses were used.

## Results

### Discovery two-sample MR analysis on the causal relationship between daytime napping and health outcomes

The study design was schematically illustrated in Figure 1. Among UKBB participants, 56.1% never/rarely took napping during daytime, 38.3% took a nap sometimes, while 5.4% took daytime naps usually and 0.2% preferred not to answer this question. The numbers of qualified IVs employed to instrument daytime napping frequency was 66 at most. Given the availability of SNPs overlapping between the exposure and outcome studies, the causal relationships between daytime napping frequency (a UKBB trait) and 1088 published non-UKBB GWAS outcomes were eventually screened in our discovery stage. We identified 19 outcomes as statistically significant outcomes causally related to daytime napping frequency.

Specifically, the daytime napping frequency was causally associated with increased risk for major depressive disorder (MDD) (p=6.8×10^−5^), migraine in bipolar disorder (p=4.5×10^−2^), class 2 obesity (p=1.3×10^−2^) and class 3 obesity (p=6.9×10^−3^), coronary heart disease (p=2.3×10^−2^), schizophrenia (p=3.2×10^−2^), and increased fasting insulin (p=1.1×10^−2^). We also observed a causal association between increased daytime napping frequency and increased levels of a number of lipid traits that are mainly related to cholesterol and cholesterol-related lipoproteins. The majority of remaining causal associations were with a wide variety of blood metabolites (Supplementary Table 1).

**Table 1.**
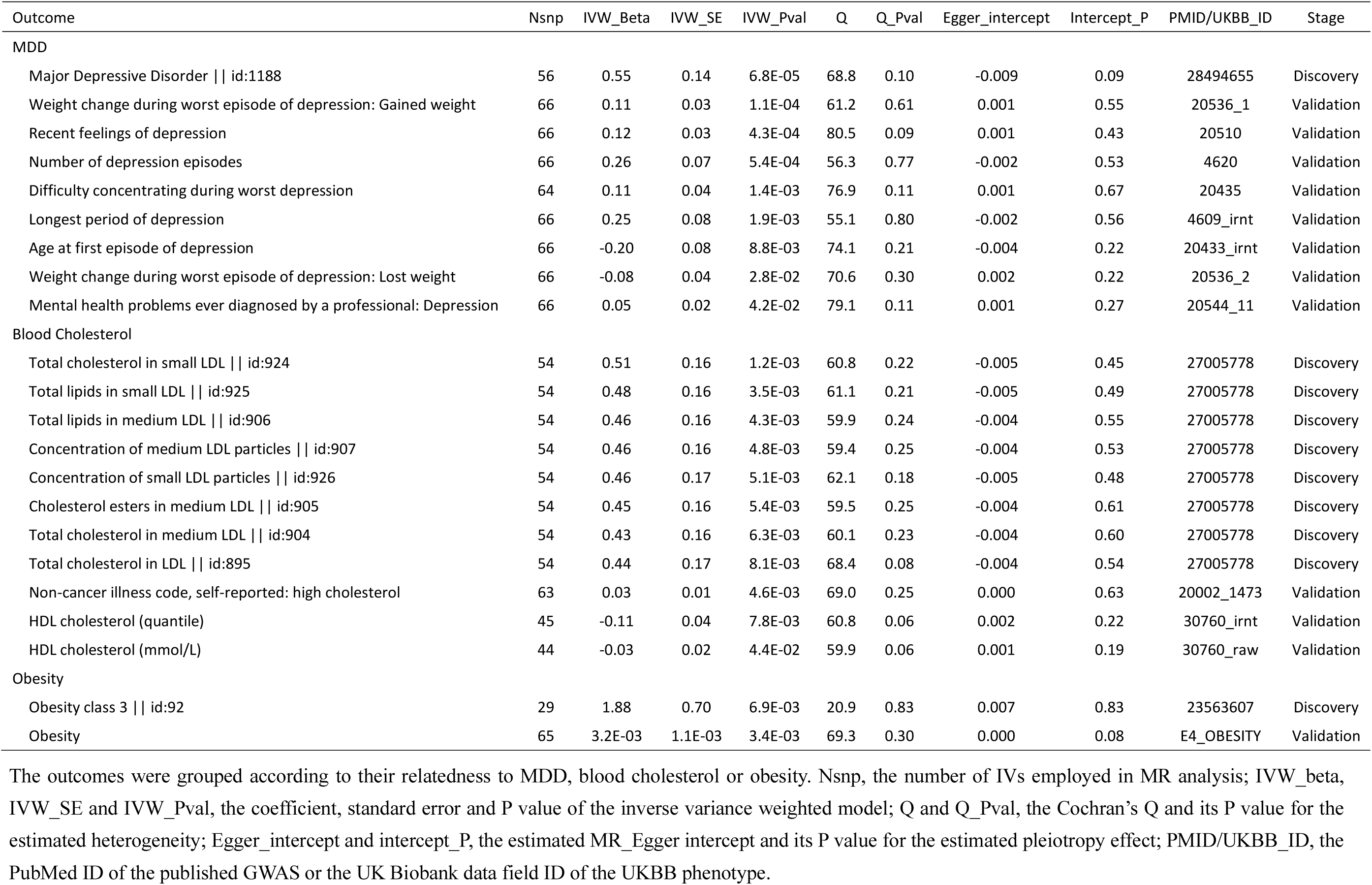
MR results of daytime napping frequency in the discovery and validation stages.

The F statistics of daytime napping frequency IVs were estimated to range from 28.1 to 1089.5. The empirical F statistic of both IV sets is greater than 10, indicating a strong instrument set. We anticipated that our discovery MR analysis had sufficient statistical power to detect a genuine causal effect (See power analysis in Supplementary Methods).

### Validation MR analysis in UK Biobank

The significant causal effects were further validated in the UKBB data (Figure 1). This was performed by using the UKBB daytime nap frequency trait as the exposure, and other UKBB traits as outcomes, therefore a one-sample MR analysis.

We observed that more frequent daytime napping is significantly associated with increased risks for depression-related traits including “weight change during worst episode of depression: Gained weight” (p=1.1×10^−4^), “recent feelings of depression” (p=4.3×10^−4^), “number of depression episodes” (p=5.4×10^−4^), “difficulty concentrating during worst depression” (p=1.4×10^−3^), “longest period of depression” (p=1.9×10^−3^), and “mental health problems ever diagnosed by a professional: Depression” (p=4.2×10^−2^). There is a causal association between more frequent napping and decreased risks for “age at first episode of depression” (p=8.8×10^−3^) as well as “weight change during worst episode of depression: Lost weight” (p=2.8×10^−2^). Frequent napping is also significantly associated with increased risk for obesity (p=3.4×10^−3^). Meanwhile, a number of lipid traits related to cholesterol were also validated where the napping frequency was associated with increased LDL cholesterol and other lipids but decreased HDL cholesterol (P<0.05 for all tests, Table 1 and Figure 2). Again, our sensitivity analysis showed no dominant effect of any single IVs in the validated causal effects.

**Figure 2.**
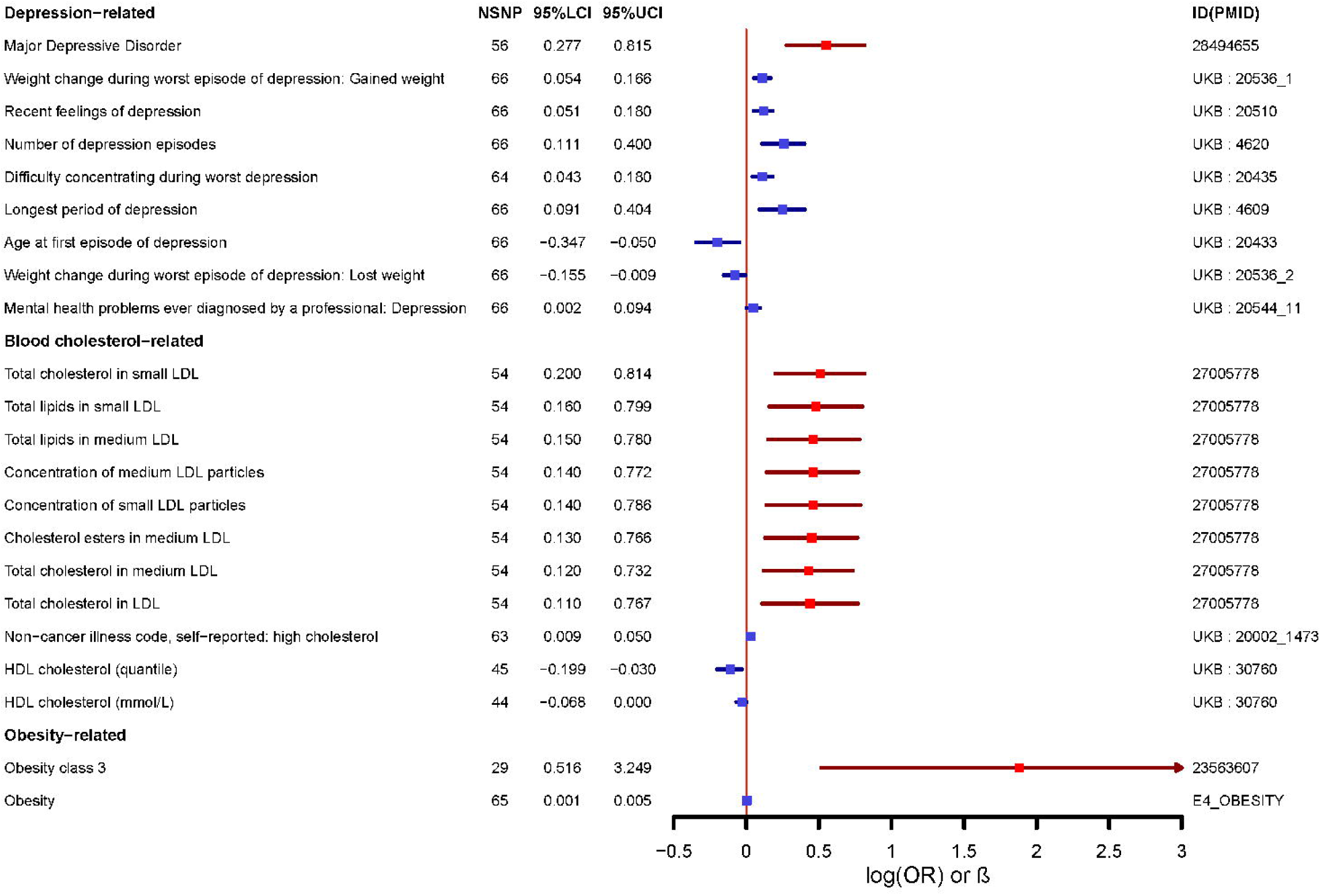
The forest plot of validated consequences of frequent daytime napping. The discovery results from non-UKBB GWAS are plotted in red, while the validation results from UKBB data are in blue.

### Reverse MR analyses

We sought to further explore whether there is a reverse causal relationship between daytime napping and the associated health outcomes. We conducted a reverse MR using MDD and obesity class 3 from non-UKBB data, as well as body mass index and obesity from UKBB data as the exposure and causally associated traits, including daytime napping frequency as the outcome. Our result showed that the exposure to MDD may result in more frequent daytime naps (IVW beta=0.035, p-value=1.8×10^−4^), while the exposure to obesity may lead to MDD, high cholesterol, lower HDL cholesterol, higher LDL cholesterol and frequent daytime napping (Supplementary Table 2).

## Discussion

Sleep is a key factor influencing both mental and physical health. With regard to the health consequences of daytime napping, the conclusions drawn from observational studies remain controversial, which is largely due to the limitations in resolving the confounding factors that were not observed in these studies. Our MR analysis for the first time provided a novel and comprehensive insight into how daytime napping habit may affect human health, by investigating its causal effect on thousands of traits simultaneously.

### Frequent daytime napping is primarily detrimental to both mental and physical health

Our discovery results indicated that daytime napping may lead to a broad spectrum of adverse consequences, including a variety of health outcomes and metabolite levels. More specifically, we found that increased napping frequency is mainly detrimental to human health, by increasing risks for depression, lipid homeostasis and obesity. These findings were especially supported by our two-stage design consisting of a discovery and a validation analysis. Our findings indicated that frequent daytime napping should be avoided especially for people who might have increased risks for depressive disorders and metabolic perturbations.

Our data suggest that taking more often nap may increase the risk of major depression, obesity as well as elevated lipid profiles. It has been long recognized that depression is closely related to obesity and deviated cardiometabolic profiles ^41-51^. Sleep patterns are also correlated with both obesity and psychopathology ^52^. To further tackle the relationships between daytime napping, obesity, and MDD, we investigated whether the exposure to obesity or MDD could result in more frequent daytime napping. Interestingly, our reverse MR analysis suggested that indeed, both obesity and MDD can cause increased nap frequency, and obesity also causally increases the risk of MDD and dysregulated lipid profiles (Supplementary Table 2). Our study thus reveals that there may be a true causation underlying these highly linked health issues, and frequent daytime napping, obesity and MDD could make a feedforward cycle to jeopardize mental and physical health.

Taken together, our study warrants continued investigations to examine whether it could improve the mental and physical health by breaking the feedforward loop between napping, obesity and depressive mood by certain intervention on napping behavior or strategy.

### Study limitations

Our study may be technically limited as well. First, the MR analysis leverages the natural randomization of alleles of genomic variants to instrument the effect of an exposure to an outcome, which mimics a randomized clinical trial, thus requires a few important assumptions especially that the instrument variants should have a direct causal relationship with the exposure without pleotropic effects that may lead to a direct impact on the outcomes. Meanwhile, the instrument variant should have effects on the outcome solely mediated by the exposure ^21^. While we have tried our best to select qualified instrument variants, our analysis cannot completely rule out the spurious pleiotropic effects using published GWAS data where the statistical power might be limited. Also, the MR analysis is preferably performed with instrument-exposure and instrument-outcome associations in nonoverlapping samples, i.e. two-sample MR ^53^. While the discovery analysis in our study is designed as a two-sample MR, given the lack of additional large instrument-nap frequency association dataset other than UKBB, our validation analysis for nap frequency is essentially a one-sample MR, thus being prone to potential bias. Also, given the large cohort of UKBB, it is still possible that some samples in the instrument-outcome associations (GWAS consortia) may partially overlap with that of UKBB. Another potential problem is that the definition of the phenotypes in the GWAS consortia studies (more clinically defined) and UKBB (medical record/self-reported) may not match well. In addition, the effect size of the instrument variants may not be sufficient. The SNP heritability of daytime napping frequency is estimated to be 8.38% in our analysis (Supplementary Methods). Thus, our MR analysis may not be able to anchor the effect of the remaining 91.62% of daytime napping frequency on health outcomes. Due to the complexity of the interactions between daytime napping and many other factors e.g. age, gender, socioeconomic status, life style, etc., it is still possible that our findings are weakened by these potential confounders. Nevertheless, we used the largest-to-date samples and datasets for this MR analysis, and the relatively high consistency between the discovery and validation analyses still generates comprehensive ever and interesting hypotheses warranting further investigations.

## Conclusions

Our study screened the causal effects of daytime napping in over 1000 human health outcomes and came up with a network of the causal effects centered by nap frequency (Figure 3). Our study concludes that frequent daytime napping may be detrimental to human physical and mental health by directly increasing the risks for obesity, MDD and high cholesterol. Our study also suggests that daytime napping could be a treatment/prevention target of both obesity and MDD. Further studies should be focused on better defined daytime napping traits (e.g. more quantitative frequency, detailed duration, napping time, etc.) in cohorts of different age groups, in particular using prospective and well-controlled designs. As such, a precision strategy for a health improvement can be eventually achieved.

**Figure 3.**
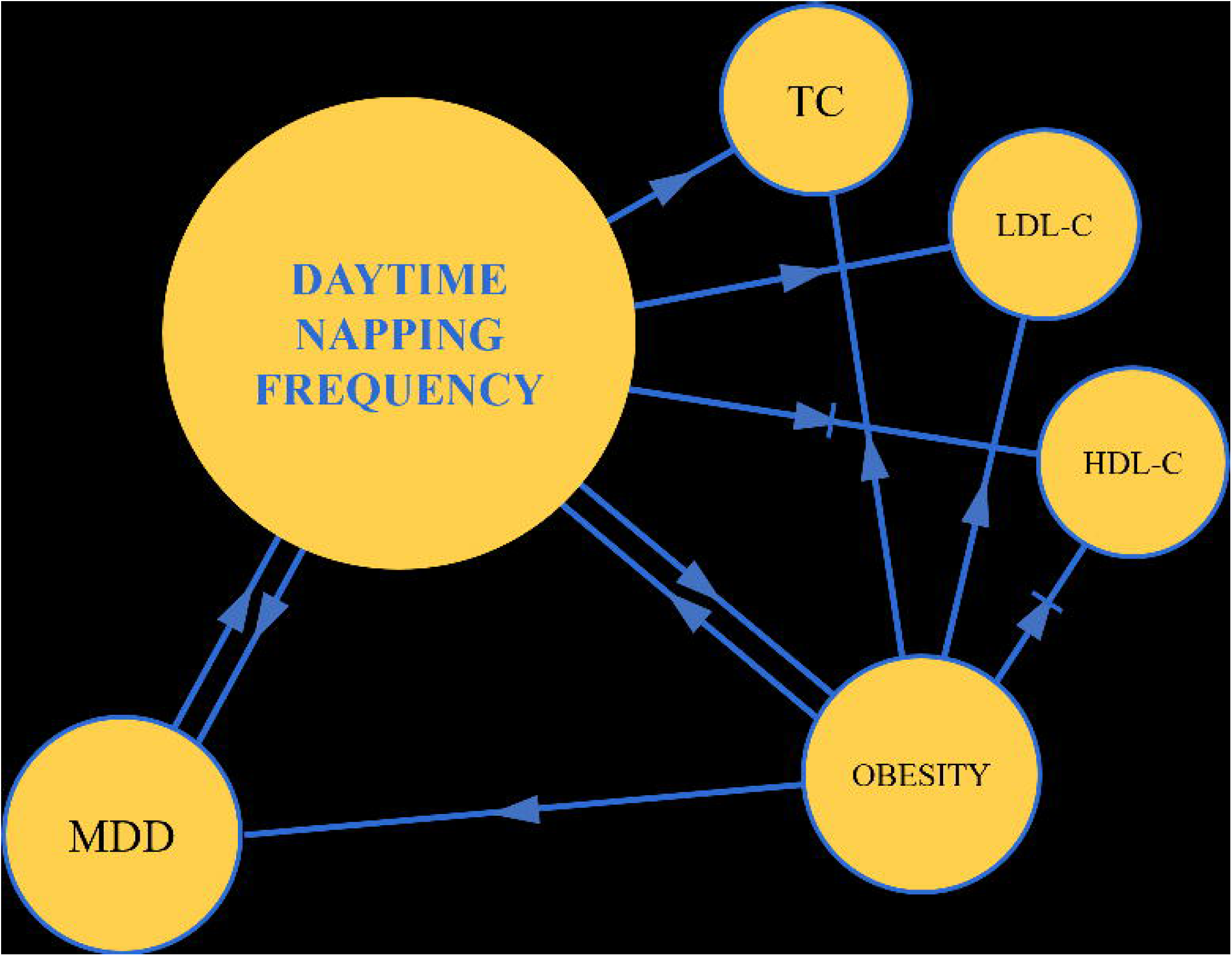
An illustrative chart of the consequences of daytime napping frequency. Lines with arrows start from an exposure and point to an outcome. An exposure increased the risk of an outcome or the level of a quantitative trait when they are connected by a sharp arrow. On the contrary, an exposure decreased the risk of an outcome or the level of a quantitative trait when they are connected by a blunt arrow.

### Summary Box

**What is already known on the health impact of daytime napping**

- The association of daytime napping and human health outcomes has been partially investigated both in observational studies and clinical trials.
- The overall and broad impact of daytime napping on human physical and mental health remained to be clarified.

**What this study adds**

- A comprehensive phenotype-wide Mendelian randomization analysis to test the causality of daytime napping on a wide variety of physical and mental health outcomes, using the largest-to-date datasets.
- Frequent daytime napping is causal to increased risks of obesity, major depressive disorder, and high cholesterol.
- Frequent daytime napping should be avoided especially in individuals with high risks for depressive and/or metabolic disorders, though a clear guideline is to be examined in future studies.

## Data Availability

Perl script and R code used in this study are available upon request from the corresponding author at. The summary statistics of published GWAS are available from MRbase webapp (http://www.mrbase.org/) and can be accessed through TwoSampleMR package (https://github.com/MRCIEU/TwoSampleMR). UKBB GWAS summary statistics are available from Benjamin Neale’s website (http://www.nealelab.is/uk-biobank, GWAS round 2).

## Acknowledgements

We thank Benjamin Neale’s group and all the genetics researchers and consortiums for making the GWAS summary data publicly available.

## Author contributions

PC and WL contributed equally to this work and are joint corresponding authors. LC and AT contributed equally to this work and are joint first authors. LC, PC, ZL and WL devised the study and finalized the data analysis plan. LC and PC collected the data. LC, AT, MZ and PC wrote the code for analysis and analyzed the data. LC, AT, ZL, XP, CH, PC and WL interpreted the results. LC, AT, PC and WL wrote the manuscript. All authors revised the manuscript and gave the permission to publish the manuscript. The corresponding authors attest that all listed authors meet authorship criteria and that no others meeting the criteria have been omitted.

## Roles of the funding sources

This work was supported in part by the start-up of the Office of Vice President for Research of Wayne State University (W.L.). P. C. was supported by Changbai Mountain Scholars Program of Jilin Provence. All sources of funding were not involved in the study design, data collection and analysis, interpretation of the results, manuscript writing, or the decision in submitting the manuscript for publication. The authors are independent from these funding sources. The authors have full access to the GWAS summary statistics of UKBB, can take the responsibility for the integrity of these summary statistics and the accuracy of the data analysis that have been done by the authors.

## Transparent Statement

The manuscript’s guarantors (LC, AT, PC, and WL) affirm that the manuscript is an honest, accurate, and transparent account of the publicly available GWAS summary statistics; that no important aspect of the study has been omitted; that any discrepancies from the study as originally planned have been explained.

## Supplementary Tables

*Supplementary Table 1. The MR results using daytime nap frequency as exposure in our discovery stage*. MRbase_ID and PMID are the ID in MRbase and pubmed database. They are used to refer to the study which reported the GWAS on the outcomes. Nsnp, number of IVs; IVW_Beta, IVW_Se and IVW_Pval, the causal effect, the standard error and the p-value of causal effect; Q_Pval, the p-value of Cochran’s Q value; Egger_intercept_P, the p-value of MR-Egger intercept. The candidate outcomes that were selected for validation are in bold.

*Supplementary Table 2. The MR result of MDD or obesity as exposure* The MR analyses with obesity class 3 and E4_obesity as exposure were done using IVs selected by exposure p-value ≤ 1 × 10^−5^. Nsnp, number of IVs; IVW_Beta, IVW_Se and IVW_Pval, the causal effect, the standard error and the p-value of causal effect; Q_Pval, the p-value of Cochran’s Q value; Egger_intercept_P, the p-value of MR-Egger intercept.

